# FLAMeS: A Robust Deep Learning Model for Automated Multiple Sclerosis Lesion Segmentation

**DOI:** 10.1101/2025.05.19.25327707

**Authors:** Emma Dereskewicz, Francesco La Rosa, Jonadab dos Santos Silva, Edward Sizer, Amit Kohli, Maxence Wynen, William A. Mullins, Pietro Maggi, Sarah Levy, Kamso Onyemeh, Batuhan Ayci, Andrew J. Solomon, Jakob Assländer, Omar Al-Louzi, Daniel S. Reich, James Sumowski, Erin S. Beck

**Affiliations:** Department of Neurology, Icahn School of Medicine at Mount Sinai, New York, NY, 10029, USA; Windreich Department of Artificial Intelligence & Human Health, Icahn School of Medicine at Mount Sinai, New York, NY 10029, USA; ICTeam, Université Catholique de Louvain, Louvain-la-Neuve, Belgium; Neuroinflammation Imaging Lab, Université Catholique de Louvain, Brussels, Belgium; Translational Neuroradiology Section, National Institute of Neurological Disorders and Stroke, National Institutes of Health, Bethesda, MD, USA; Cliniques Universitaires Saint-Luc, Université Catholique de Louvain, Brussels, Belgium; Department of Neurology, University of Vermont, Burlington, VT; Center for Biomedical Imaging, Department of Radiology, New York University School of Medicine, New York, NY, USA; Cedars-Sinai Medical Center, Los Angeles, CA, USA

## Abstract

**Background and Purpose:** Assessment of brain lesions on MRI is crucial for research in multiple sclerosis (MS). Manual segmentation is time consuming and inconsistent. We aimed to develop an automated MS lesion segmentation algorithm for T2-weighted fluid-attenuated inversion recovery (FLAIR) MRI.

**Methods:** We developed FLAIR Lesion Analysis in Multiple Sclerosis (FLAMeS), a deep learning-based MS lesion segmentation algorithm based on the nnU-Net 3D full-resolution U-Net and trained on 668 FLAIR 1.5 and 3 tesla scans from persons with MS. FLAMeS was evaluated on three external datasets: MSSEG-2 (n=14), MSLesSeg (n=51), and a clinical cohort (n=10), and compared to SAMSEG, LST-LPA, and LST-AI. Performance was assessed qualitatively by two blinded experts and quantitatively by comparing automated and ground truth lesion masks using standard segmentation metrics.

**Results:** In a blinded qualitative review of 20 scans, both raters selected FLAMeS as the most accurate segmentation in 15 cases, with one rater favoring FLAMeS in two additional cases. Across all testing datasets, FLAMeS achieved a mean Dice score of 0.74, a true positive rate of 0.84, and an F1 score of 0.78, consistently outperforming the benchmark methods. For other metrics, including positive predictive value, relative volume difference, and false positive rate, FLAMeS performed similarly or better than benchmark methods. Most lesions missed by FLAMeS were smaller than 10 mm³, whereas the benchmark methods missed larger lesions in addition to smaller ones.

**Conclusions:** FLAMeS is an accurate, robust method for MS lesion segmentation that outperforms other publicly available methods.

## 1 Introduction

Multiple sclerosis (MS) is a chronic immune-mediated condition of the central nervous system (CNS), characterized by inflammatory demyelination and neurodegeneration^1^. Inflammatory demyelination results in focal brain lesions that are hyperintense on T2-weighted MRI images. Clinically, these lesions are key for MS diagnosis^2^, assessment of response to treatment^3^, and predictions of future disability^4^. Assessment of lesion number and volume is also essential for nearly all MS imaging research. Reliable lesion segmentation provides the foundation for studying advanced imaging biomarkers, including central vein sign and paramagnetic rim lesions^5^.

Manual segmentation of MS lesions is time consuming and susceptible to interrater variability, particularly in people with high lesion burden. Automated methods offer faster and more consistent results, making them a powerful tool for MS research. Among these, convolutional neural network-based approaches have emerged as the state-of-the-art technique^6–9^, consistently outperforming traditional machine learning methods and achieving the best performance in MS lesion segmentation challenges^10–12^.

The vast majority of automated lesion segmentation methods require both a T2-weighted (w) FLAIR and a T1w image as input. These algorithms are typically trained on high-resolution, research-grade scans. However, since these scans are not routinely acquired in clinical practice, their applicability is often limited to research settings. Additionally, since the two image types must be co-registered before processing, any misalignment or registration errors can introduce inaccuracies in the lesion segmentation, further complicating their use in diverse, multi-center research datasets and real-world clinical settings.

In this study, we aimed to develop an accurate and effective automated method for MS lesion segmentation from T2w FLAIR images alone that improves upon currently available algorithms. Our proposed algorithm, FLAIR Lesion Analysis in Multiple Sclerosis (FLAMeS), is a deep learning-based segmentation method that is trained on a diverse dataset of 668 scans acquired with 1.5 and 3 tesla (T) MRI scanners. We evaluate the performance of FLAMeS in comparison to two established and publicly available algorithms, SAMSEG and LST. As testing datasets, we consider both research- and clinical-grade MRI scans acquired on several different MRI scanners from four different manufacturers at 1.5 and 3T.

## 2 Methods

### 2.1 Datasets

All datasets used in this study were stripped of protected health information. For all private datasets, participants provided informed consent as part of an IRB-approved research protocol.

#### Training Set

The training set was composed of 668 T2-weighted FLAIR brain MRI scans from 575 people with MS (pwMS), imaged at seven sites. The training set included images from three publicly and five privately available datasets, acquired at 1.5 and 3T. Ground truth lesion masks were either manually segmented by experts^12–14^ (n=97) or derived from other automated lesion segmentation tools^15–17^ and subsequently refined through manual correction (n=571). The median number of lesions per scan was 49 (IQR: 27–92; range 0–426), and the median total lesion volume was 4574 mm^3^ (IQR: 1429.0–12804.2 mm^3^; range 0.0–88930.0 mm^3^).

We initially trained the FLAMeS model using 161 FLAIR scans. To enhance performance, we later expanded the training dataset by incorporating an additional 507 scans from three external datasets^16^. The lesion masks for 307 of these additional training scans were initially generated by FLAMeS and then manually refined by a human rater.

#### Testing Sets

##### MSSEG-2 challenge

The test set from the MSSEG-2 challenge^10^ dataset (n=14) from diverse sites and scanner manufacturers and models was used for testing (Table 1, demographic data not available). Lesions were manually segmented on 3D FLAIR images using ITK Snap^18^ by E.D., followed by review and adjustment if needed by E.S.B. The 14 cases were selected from the larger MSSEG-2 challenge set, with the goal of including images acquired across various 3T MRI scanners with differing manufacturers and models (Table 1).

**Table 1.**
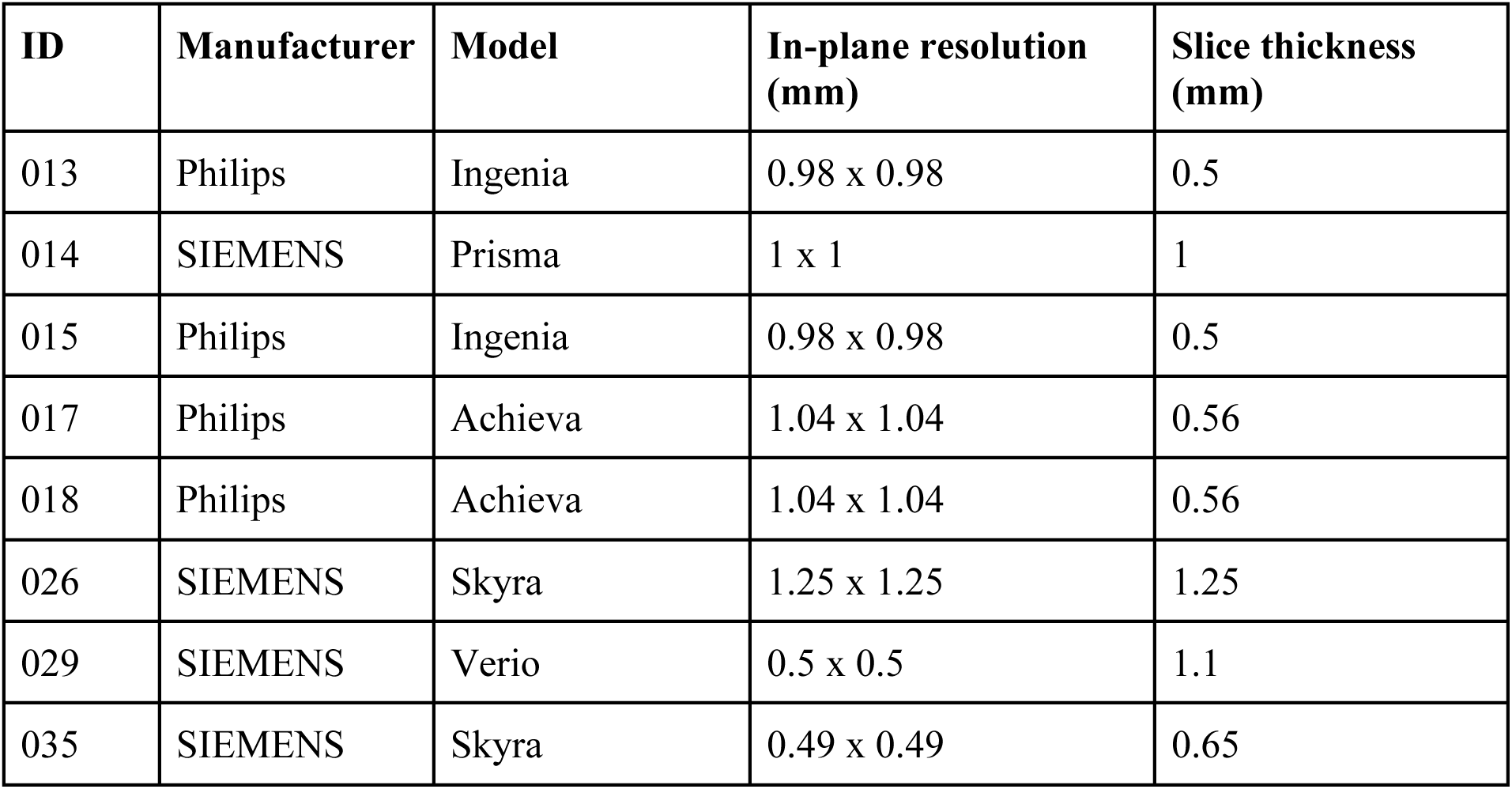
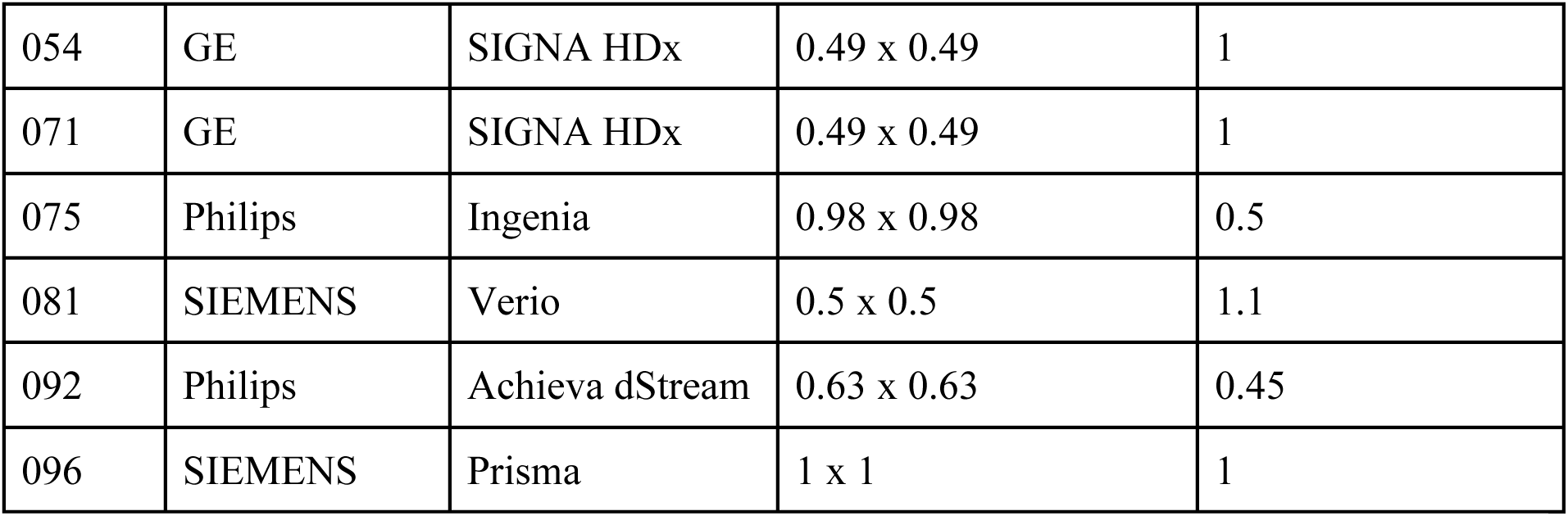
MSSEG-2 testing dataset (n=14).

| ID | Manufacturer | Model | In-plane resolution (mm) | Slice thickness (mm) |
| --- | --- | --- | --- | --- |
| 013 | Philips | Ingenia | 0.98 x 0.98 | 0.5 |
| 014 | SIEMENS | Prisma | 1 x 1 | 1 |
| 015 | Philips | Ingenia | 0.98 x 0.98 | 0.5 |
| 017 | Philips | Achieva | 1.04 x 1.04 | 0.56 |
| 018 | Philips | Achieva | 1.04 x 1.04 | 0.56 |
| 026 | SIEMENS | Skyra | 1.25 x 1.25 | 1.25 |
| 029 | SIEMENS | Verio | 0.5 x 0.5 | 1.1 |
| 035 | SIEMENS | Skyra | 0.49 x 0.49 | 0.65 |
| 054 | GE | SIGNA HDx | 0.49 x 0.49 | 1 |
| 071 | GE | SIGNA HDx | 0.49 x 0.49 | 1 |
| 075 | Philips | Ingenia | 0.98 x 0.98 | 0.5 |
| 081 | SIEMENS | Verio | 0.5 x 0.5 | 1.1 |
| 092 | Philips | Achieva dStream | 0.63 x 0.63 | 0.45 |
| 096 | SIEMENS | Prisma | 1 x 1 | 1 |

##### MSLesSeg challenge

The MSLesSeg challenge was held at the International Conference on Pattern Recognition (ICPR) in 2024^11,19,20^. It included a training dataset of images from 53 pwMS imaged clinically at 1.5 T at different hospital centers in Catania, Italy. The imaging protocols varied, but they all included FLAIR and T1-w images. Ground truth MS lesion annotations were performed using Jim 9 (Xinapse Systems, UK), a semi-automated segmentation tool, refined manually by two raters and reviewed by an experienced neurologist specialized in MS. The lesion masks were publicly released as part of the challenge dataset. The images were released publicly after the following pre-processing steps: de-identification, conversion to NIfTI format, linear registration to the standard MNI152 template space in 1 × 1 × 1 mm resolution, and skull-stripping. Original scan parameters for this testing set, including original resolution, were not available. For our analysis, we excluded two cases from the MSLesSeg challenge due to failed skull-stripping, leading to errors by all automated segmentation methods. The demographics of the pwMS included are not known.

##### Clinical dataset

To evaluate the performance of the segmentation methods on diverse clinical data, we selected 10 clinical-grade FLAIR scans of pwMS (8 [80%] female, mean age 46 ± 12 years, mean time since diagnosis 9 ± 8 years, 9 RRMS, 1 SPMS) performed at Mount Sinai. These images varied in manufacturer, model, field strength, slice thickness, and dimensions to best represent the diversity in clinical MRI acquisition (Table 2). MS lesions were manually segmented on ITK Snap^18^ by E.D. and subsequently reviewed and adjusted as needed by E.S.

**Table 2.** Clinical testing dataset (n=10). *Reconstructed slice thickness (acquired slice thickness, if different).

| ID | Manufacturer | Model | Software version | Field strength (T) | In-plane resolution (mm) | Slice thickness *(mm) | Acquisition |
| --- | --- | --- | --- | --- | --- | --- | --- |
| 1 | GE | Signa HDxt | 23 LX | 1.5 | 0.45 x 0.45 | 0.9 (1.8) | 3D |
| 2 | GE | Discover y MR750 | 25 LX | 3 | 0.47 x 0.47 | 0.8 (1.6) | 3D |
| 3 | SIEMENS | Skyra | Syngo MR E11 | 3 | 0.49 x 0.49 | 1 | 3D |
| 4 | GE | Signa Excite | 11 LX | 1.5 | 0.43 x 0.43 | 6 | 2D - Ax |
| 5 | SIEMENS | Skyra | Syngo MR E11 | 3 | 0.49 x 0.49 | 3 | 2D - Ax |
| 6 | SIEMENS | Aera | Syngo MR E11 | 1.5 | 0.69 x 0.69 | 5 | 2D - Ax |
| 7 | GE | Signa HDxt | 23 LX | 1.5 | 0.43 x 0.43 | 0.8 (1.6) | 3D |
| 8 | SIEMENS | Aera | Syngo MR E11 | 1.5 | 0.86 x 0.86 | 5.5 | 2D - Ax |
| 9 | SIEMENS | Aera | Syngo MR E11 | 1.5 | 0.98 x 0.98 | 1 | 3D |
| 10 | SIEMENS | Verio | Syngo MR B19 | 3 | 0.69 x 0.69 | 5 | 2D - Ax |

### 2.2 FLAMeS

FLAMeS is based on the nnU-Net 3D full-resolution U-Net architecture^21^, optimized for segmenting MS lesions from FLAIR images. The model takes as input 3D volumetric data (FLAIR images), leveraging a six-stage U-Net architecture with progressively increasing feature channels: 32, 64, 128, 256, 320, and 320. Each stage consists of two convolutional layers with 3 × 3 × 3 kernels, followed by instance normalization and LeakyReLU activation. Downsampling is performed via strided convolutions, with strides of (2,2,2) at most levels, except for the first stage, which maintains full resolution, and the final stage, which has an asymmetric stride of (1,2,2) because of the anisotropic voxel spacing. The decoder follows a symmetric structure with transposed convolutions for upsampling. FLAMeS is trained for 8,000 epochs with a batch size of 2, using stochastic gradient descent (SGD) with momentum (0.99) and weight decay (3e-5). The standard nnUNet loss function is a combination of cross-entropy and dice loss.

We did an ablation study comparing the standard loss function with a loss function that combines Tversky loss and Focal loss, balancing sensitivity and specificity while addressing the class imbalance typical in lesion segmentation tasks. The Tversky loss parameters were α = 0.55 and β = 0.45, adjusting the trade-off between false positives and false negatives, where slightly higher α values penalize false positives more heavily. The Focal loss used γ = 1.5 to focus learning on harder-to-classify lesions, with an additional weighting factor α = 0.20 to control the contribution of positive samples. To improve generalizability, nnU-Net’s built-in deep supervision strategy is employed, enforcing loss constraints at multiple feature levels. The model ensemble combines predictions from five independently trained networks, further enhancing robustness across diverse MRI datasets.

FLAMeS follows the nnU-Net standardized preprocessing pipeline to ensure robustness across different datasets and scanner manufacturers. All input FLAIR images are resampled to a uniform voxel spacing of 1.0 × 0.9 × 0.9 mm, using third-order spline interpolation for intensity data and first-order interpolation for segmentation masks. Z-score intensity normalization is applied within the brain mask to standardize signal distributions across scans. To minimize unnecessary background and optimize GPU memory usage, images are cropped around the brain region. During training, nnU-Net’s standard data augmentation strategies — including intensity scaling, rotation, flipping, and elastic deformations — are used to reduce the risk of overfitting.

The only pre-processing step needed for FLAMeS is skull-stripping, which we performed with the SynthSeg^22^ tool from Freesurfer^23^, selecting the *-no-csf* flag, which excludes the cerebrospinal fluid (CSF) from the skull-stripped image.

We developed a web app based on the Hugging Face Spaces platform^24^ (https://huggingface.co/spaces/FrancescoLR/FLAMeS) to publicly deploy FLAMeS, enabling users to upload skull-stripped FLAIR images and download automated lesion segmentations. Built using Python and Gradio library, the web app features an intuitive user interface, real-time processing status updates, and downloadable output files, including the binary lesion mask, a mask with individual labels for each lesion, and a visual overlay of the segmentation on the input image. This deployment ensures accessibility for researchers and clinicians, facilitating seamless integration of FLAMeS into various neuroimaging research workflows without requiring local computational resources. Inference through the web app takes approximately one minute per input image. For accelerated performance and large-scale processing, we recommend running FLAMeS in a GPU-enabled environment.

### 2.3 Evaluation

#### Benchmark methods

● **LST:** The Lesion Segmentation Toolbox (LST)^25^ provides three distinct methods for MS lesion segmentation: LST-LGA (Lesion Growth Algorithm), LST-LPA (Lesion Prediction Algorithm), and LST-AI (Artificial Intelligence-based segmentation)^7^. Briefly, LST-LPA is a binary regression model that integrates the lesion belief map with predefined parameters. These parameters were initially derived using logistic regression during the tool’s development to compute the lesion probability map. The final binary lesion mask is then obtained by applying a threshold of 0.5 to this probability map. The most recently described LST-AI is a deep learning-based extension of LST that consists of an ensemble of three 3D U-Nets architectures. The networks were trained on 491 pairs of T1w and FLAIR images, collected in-house from a 3T MRI scanner, and their respective manual lesion segmentations. A combination of binary cross-entropy and Tversky loss was used to increase the sensitivity to the heterogeneous MS lesions. Following the authors’ recommendations, we ran LST-LPA on the datasets where only a FLAIR image was available, while LST-AI was used when both FLAIR and T1w scans were available.
● **SAMSEG:** The Sequence Adaptive Multimodal SEGmentation (SAMSEG) algorithm^26^ is part of Freesurfer^23^ and can operate with a single MRI contrast but also supports multiple modalities. The algorithm was trained on manual segmentations from 212 MS subjects. SAMSEG employs a deformable probabilistic atlas as a segmentation prior, which is iteratively adapted to the input data. Each voxel is assigned to the most probable brain structure, including lesions. The final binary lesion mask is generated by including voxels classified as lesions while assigning a value of zero to all remaining ones. In this study, FLAIR images were always provided as input, and when available, T1w images were included to enhance segmentation accuracy.

#### Blinded qualitative evaluation

We selected 20 FLAIR scans, including the 10 clinical testing set cases and 10 randomly selected from the MSLesSeg test set. Each scan was paired with lesion masks generated by FLAMeS, LST-AI, and SAMSEG. Two independent raters, blinded to the method used, evaluated all three segmentations side by side for each scan. Segmentations were rated on a 4-point scale. A score of 1 reflected high accuracy with little to no need for adjustment, 2 indicated minor issues such as a few small missed lesions, 3 denoted multiple missed lesions or boundary inaccuracies, and 4 represented substantial errors, including large missed or poorly segmented lesions. Each rater also indicated the segmentation that they deemed most accurate for each case.

#### Metrics

To quantitatively evaluate the segmentation tools, we computed segmentation metrics using an in-house Python script. Lesions were defined as clusters of connected voxels using 3D 26-connectivity. For voxel-wise metrics, we considered the Dice similarity coefficient (DSC), the normalized DSC (nDSC)^27^, the positive predictive value (PPV), and the relative volume difference (RVD). They are defined as follows:

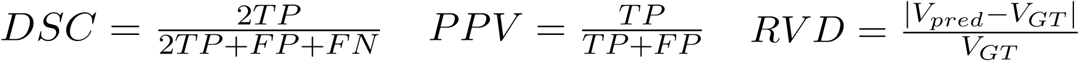

where TP (true positive) is the number of correctly predicted lesion voxels, FP (false positive) is the number of non-lesion voxels incorrectly classified as lesion, FN (false negative) is the number of lesion voxels missed by the automated tool, V_pred_ is the total lesion volume in the predicted segmentation, and V_GT_ is the total lesion volume in the ground truth segmentation.

We also assess lesion-wise segmentation performance using the Lesion True Positive Rate (LTPR), Lesion False Positive Rate (LFPR), and the Lesion-wise F1 Score (F1). These metrics evaluate the model’s ability to accurately detect individual lesions, as opposed to voxel-level segmentation accuracy. The definitions of these metrics are as follows:

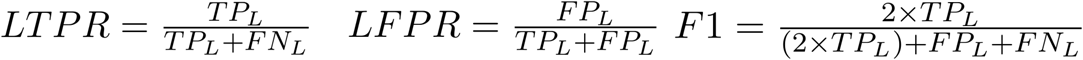

where TP_L_ (true positive lesions) is the number of correctly detected lesions, FN_L_ (false negative lesions) is the number of true lesions missed by the tool, and FP_L_ (false positive lesions) is the number of incorrectly predicted lesions.

#### Statistical analysis

Given the relatively small sample size, the presence of outliers, and strong evidence of non-normality, all pairwise comparisons were performed using the Wilcoxon signed-rank test with median values reported. We accounted for multiple comparisons using the Benjamini-Hochberg correction for false discovery rate. All significant results remained statistically significant after adjustment. The data were analyzed using IBM SPSS Statistics (Version 30.0. Armonk, NY: IBM Corp).

## 3 Results

### 3.1 Qualitative Evaluation

Lesion segmentation performance was evaluated on three testing datasets through both qualitative and quantitative comparisons with ground truth masks. Visual inspection showed that FLAMeS-generated masks more closely matched the ground truth than those from benchmark methods, with more precise lesion boundaries and fewer visible false positives or missed lesions (Figure 1). In a blinded qualitative assessment in which two raters evaluated FLAMeS, SAMSEG, and LST-AI masks side by side, both raters chose FLAMeS as the most accurate segmentation in 15 out of 20 cases. In two additional cases, one rater chose FLAMeS while the other preferred a benchmark method, and in the remaining three cases, both raters preferred a benchmark method. When evaluating the amount of manual correction needed (rated on a scale from 1 to 4, where 1 indicates minimal adjustment and 4 indicates extensive adjustment), rater 1 assigned FLAMeS an average score of 1.7, compared to 3.1 for SAMSEG and 2.5 for LST-AI. Rater 2 gave FLAMeS an average score of 2.3, versus 3.4 for SAMSEG and 3.1 for LST-AI.

**Figure 1.**
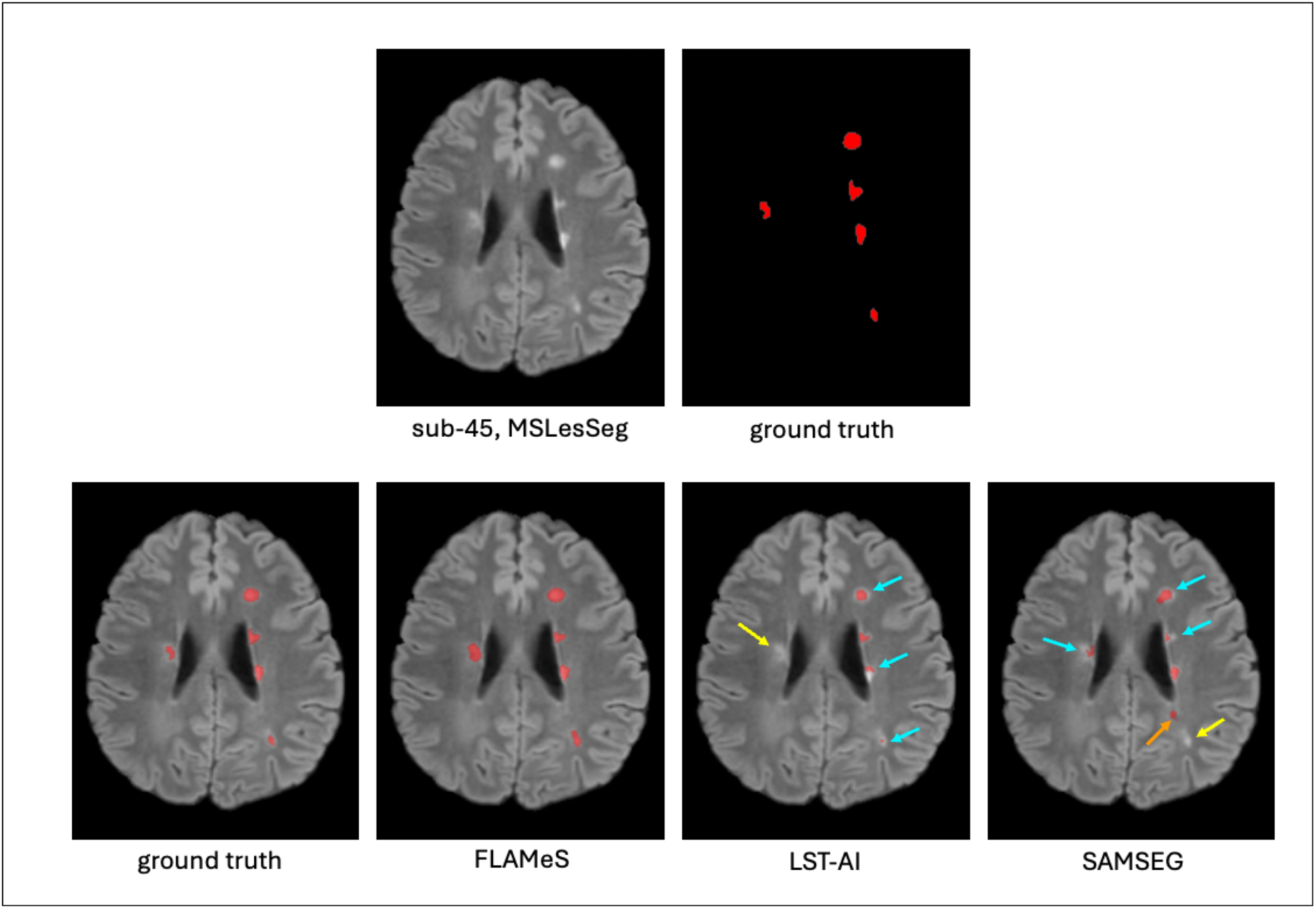
FLAMeS generates qualitatively superior lesion masks. The top row shows an axial FLAIR slice and the ground truth lesion mask. The bottom row displays the lesion masks from each method overlaid on the FLAIR image. Multiple lesions are missed (yellow arrows), undersegmented (blue arrows), or falsely segmented (orange arrow) by LST-AI and SAMSEG.

FLAMeS exhibits a slight tendency toward oversegmentation, particularly in regions with intermediate signal intensity between focal lesions and normal-appearing white matter, often with ill-defined borders. This behavior is also observed in benchmark methods (Figure 2A). To attempt to address this limitation, we conducted an ablation study comparing the standard nnUNet loss function with a balanced combination of Tversky and Focal losses. However, visual assessment of the resulting segmentations revealed minimal differences.

**Figure 2.**
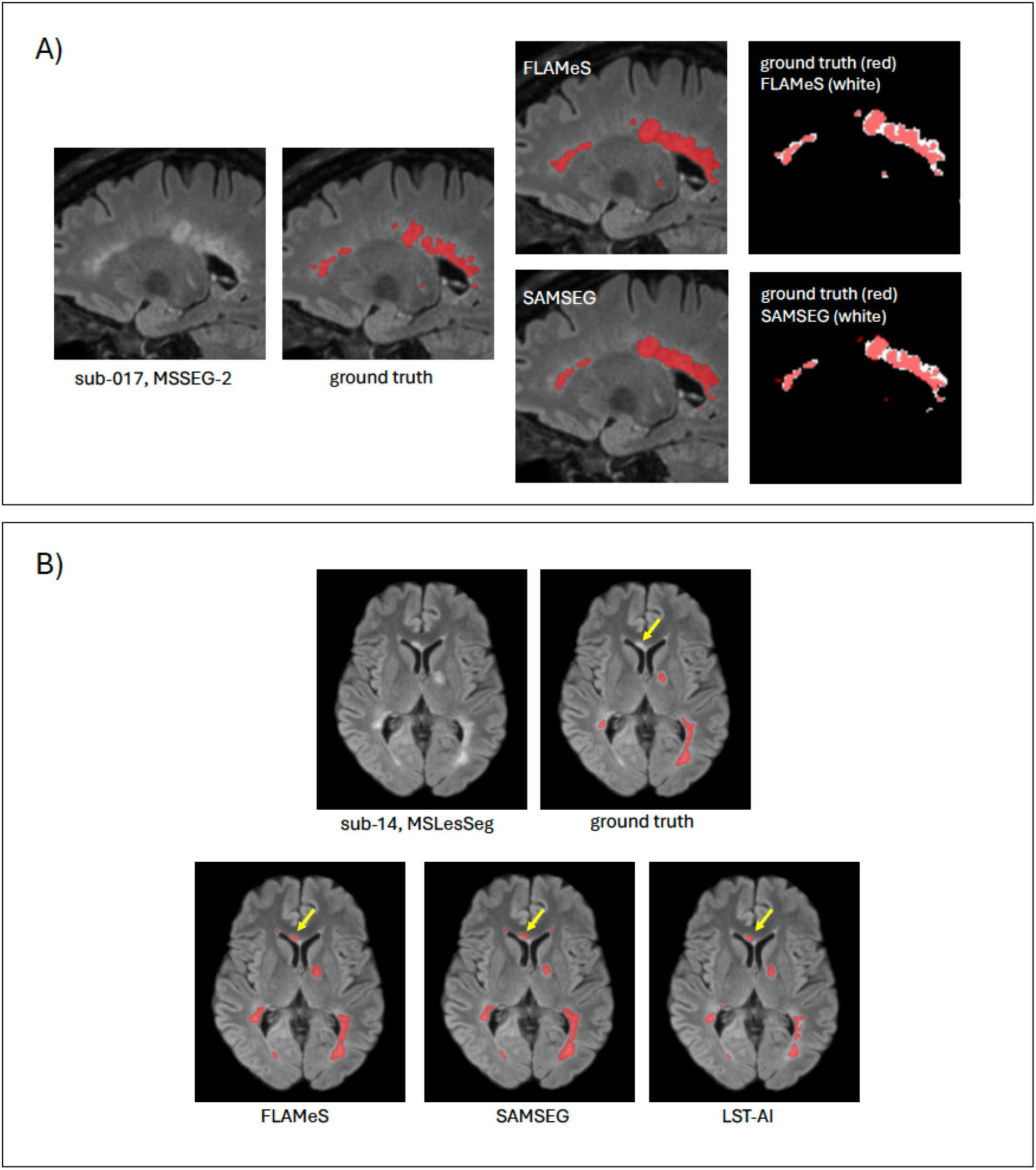
Assessment of false-positive voxels and apparently false-positive lesions identified by automated methods. (A) Oversegmentation of diffusely abnormal white matter surrounding lesions by FLAMeS and SAMSEG. (B) Example of a periventricular hyperintensity segmented by all three automated methods but not present on the MSLesSeg ground truth mask (yellow arrow). On manual review, this area was judged to be a missed lesion on the original ground truth masks.

Additionally, our review of the MSLesSeg masks revealed numerous small lesions that our raters would have annotated but were missing from the ground truth masks and were thus classified as false positives by our metrics script (Figure 2B).

### 3.2 Quantitative Evaluation

#### MSSEG-2

In the MSSEG-2 testing set, FLAMeS outperforms benchmark methods across all voxel-wise and lesion-wise metrics, except for LFPR, which was similar to that of SAMSEG but better than LST-LPA (Table 3). FLAMeS correctly identifies 70% of reference lesions — significantly more than LST-LPA (37%, p<0.001) and SAMSEG (13%, p<0.001) — while maintaining a low false positive rate of just 5%. Additionally, FLAMeS achieves an F1 score of 0.81, substantially higher than LST-LPA (0.37, p<0.001) and SAMSEG (0.13, p<0.001), demonstrating a superior balance of precision and recall. FLAMeS also produces a significantly lower relative volume difference (RVD) than the benchmark methods, with an 18% discrepancy between the automated and manual lesion masks. In comparison, RVD was 55% for SAMSEG (p=0.003), and 44% for LST-LPA (p=0.03).

**Table 3.** Performance comparison of segmentation methods on the MSSEG-2 testing dataset (n=14). Median values are reported for each metric. Pairwise comparisons were performed using the Wilcoxon signed-rank test. Values in bold indicate best performance for each metric. Abbreviations: DSC, Dice Similarity Coefficient; nDSC, normalized Dice Similarity Coefficient; PPV, Positive Predictive Value; RVD, Relative Volume Difference; LTPR, Lesion True Positive Rate; LFPR, Lesion False Positive Rate. Asterisks indicate comparisons between FLAMeS and LST, * p<0.05, ** p<0.01, *** p<0.001. Daggers indicate comparisons between FLAMeS and SAMSEG, † p<0.05, †† p<0.01, †† p<0.001

| Tool | voxel-wise |  |  |  | lesion-wise |  |  |
| --- | --- | --- | --- | --- | --- | --- | --- |
|  | DSC | nDSC | PPV | RVD | LTPR | LFPR | F1 |
| FLAMeS | **0.72**<sup>***†††</sup> | **0.75**<sup>**††</sup> | **0.96**<sup>***†</sup> | **0.18**<sup>*††</sup> | **0.70**<sup>***†††</sup> | 0.05<sup>***</sup> | **0.81**<sup>***†††</sup> |
| LST-LPA | 0.51 | 0.68 | 0.50 | 0.48 | 0.37 | 0.35 | 0.38 |
| SAMSEG | 0.47 | 0.66 | 0.93 | 0.55 | 0.13 | **0.04** | 0.23 |

#### MSLesSeg

In the MSLesSeg testing dataset, FLAMeS significantly outperformed LST-AI across all voxel-wise and lesion-wise metrics (p<0.05 or less for all comparisons) (Table 4, Figure 3). FLAMeS also outperformed SAMSEG on most metrics. While no significant differences were observed between FLAMeS and SAMSEG for PPV (p=0.05) or RVD (p=0.49), SAMSEG had a lower LFPR than FLAMeS (0.21 vs. 0.26, p=0.007). Notably, FLAMeS achieved a significantly higher LTPR (0.86) than LST-AI (0.71, p<0.001) and SAMSEG (0.43, p<0.001). It also demonstrated a significantly higher F1 score (0.77) than both LST-AI (0.57, p<0.001) and SAMSEG (0.53, p<0.001) (Table 4).

**Figure 3.**
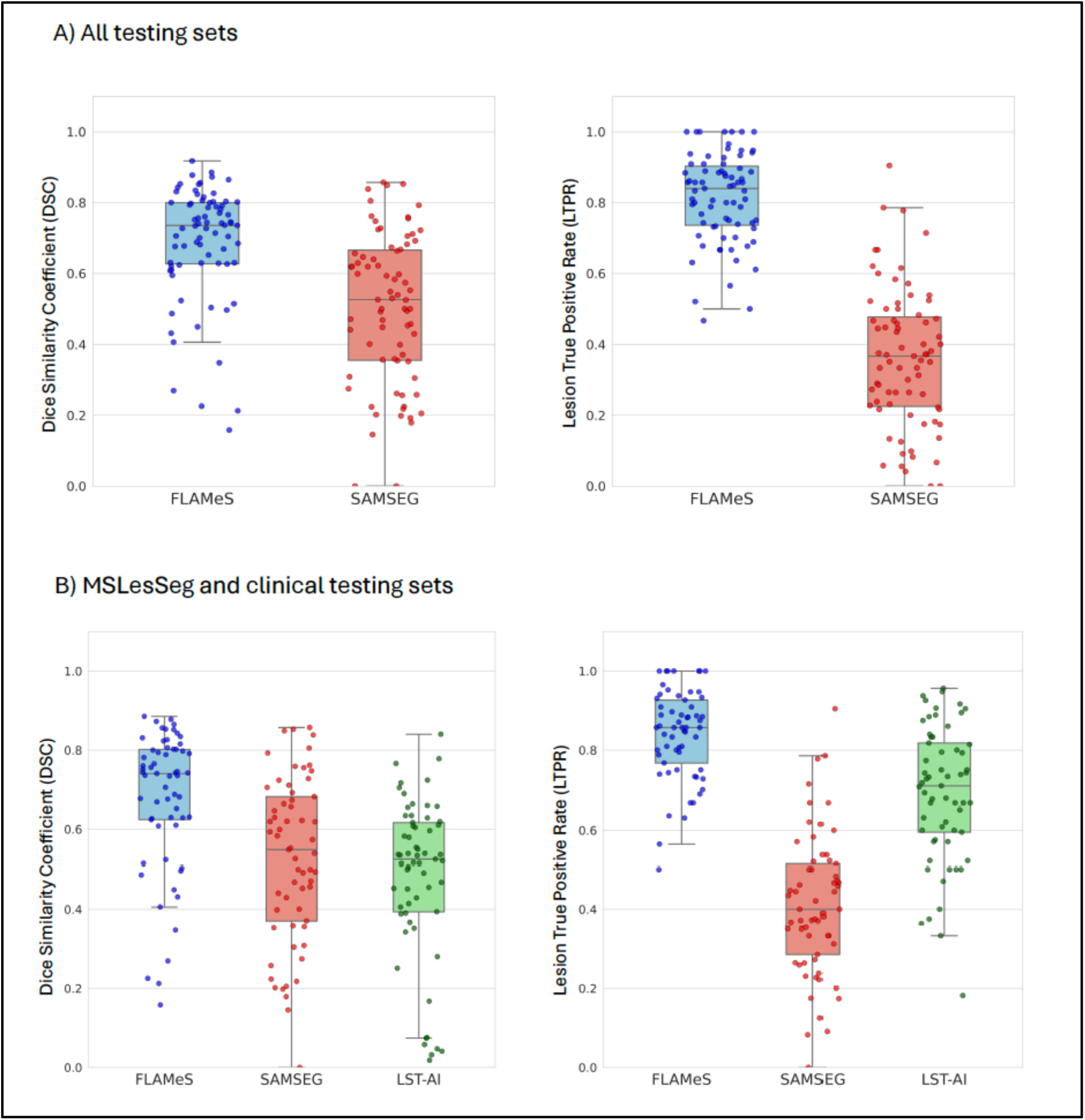
FLAMeS outperforms SAMSEG and LST-AI in DSC and LTPR. Box plots display the median (center line), interquartile range (IQR, box edges), and whiskers extending up to 1.5 × IQR.

**Table 4.** Performance comparison of segmentation methods on the MSLesSeg testing dataset (n=51). Median values are reported for each metric. Pairwise comparisons were performed using the Wilcoxon signed-rank test. Values in bold indicate best performance for each metric. Abbreviations: DSC, Dice Similarity Coefficient; nDSC, normalized Dice Similarity Coefficient; PPV, Positive Predictive Value; RVD, Relative Volume Difference; LTPR, Lesion True Positive Rate; LFPR, Lesion False Positive Rate. Asterisks indicate comparisons between FLAMeS and LST, *p<0.05, ** p<0.01, *** p<0.001. Daggers indicate comparisons between FLAMeS and SAMSEG, † p<0.05, †† p<0.01, †† p<0.001

| Tool | voxel-wise |  |  |  | lesion-wise |  |  |
| --- | --- | --- | --- | --- | --- | --- | --- |
|  | DSC | nDSC | PPV | RVD | LTPR | LFPR | F1 |
| FLAMeS | **0.74**<sup>***†††</sup> | **0.70**<sup>***†††</sup> | 0.72<sup>***</sup> | **0.17**<sup>*</sup> | **0.87**<sup>***†††</sup> | 0.26<sup>***††</sup> | **0.77**<sup>***†††</sup> |
| LST-AI | 0.43 | 0.52 | 0.45 | 0.37 | 0.71 | 0.44 | 0.57 |
| SAMSEG | 0.55 | 0.59 | **0.75** | 0.31 | 0.43 | **0.21** | 0.53 |

#### Clinical

In the clinical testing set, the proposed method outperforms the benchmark models across all metrics except for nDSC, where LST-AI matches FLAMeS with a score of 0.75 (Table 5, Figure 3). FLAMeS achieves a higher DSC of 0.68 compared to 0.63 for LST-AI (p=0.009) and 0.51 for SAMSEG (p=0.005). FLAMeS’ LFPR is 0.03, lower than LST-AI (0.24, p=0.02) but similar to SAMSEG (0.11, p=0.89). Additionally, FLAMeS attains an F1 score of 0.78, compared to 0.74 for LST-AI (p=0.009) and 0.50 for SAMSEG (p=0.007).

**Table 5.**
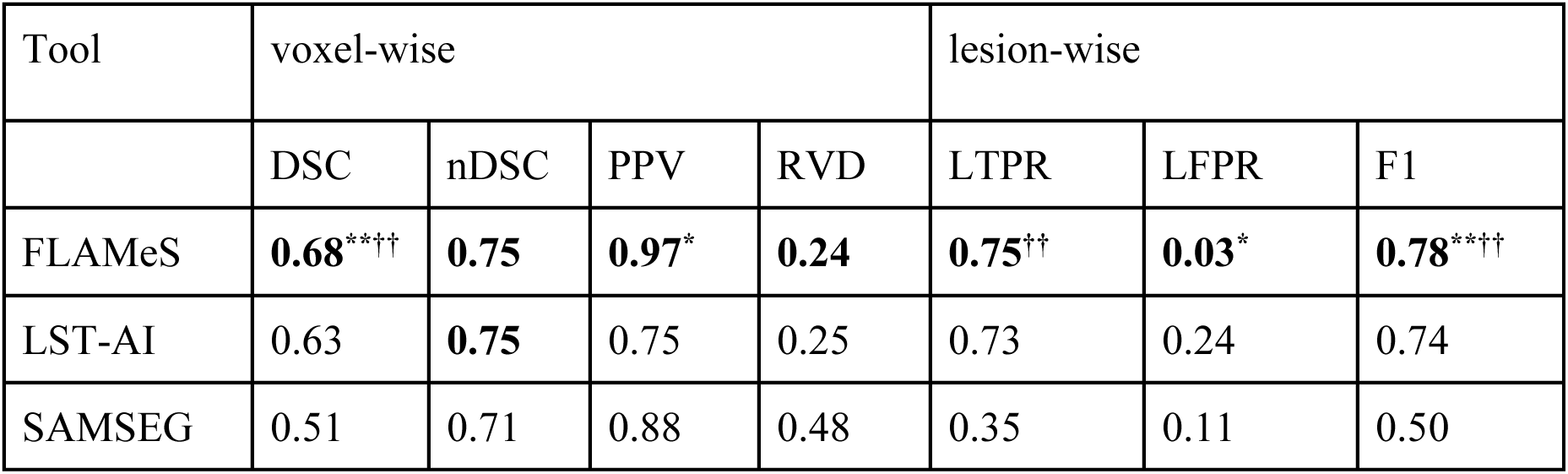
Performance comparison of segmentation methods on the clinical testing dataset (n=10). Median values are reported for each metric. Pairwise comparisons were performed using the Wilcoxon signed-rank test. Values in bold indicate best performance for each metric. Abbreviations: DSC, Dice Similarity Coefficient; nDSC, normalized Dice Similarity Coefficient; PPV, Positive Predictive Value; RVD, Relative Volume Difference; LTPR, Lesion True Positive Rate; LFPR, Lesion False Positive Rate. Asterisks indicate comparisons between FLAMeS and LST, *p<0.05, ** p<0.01, *** p<0.001. Daggers indicate comparisons between FLAMeS and SAMSEG, † p<0.05, †† p<0.01, †† p<0.001

| Tool | voxel-wise |  |  |  | lesion-wise |  |  |
| --- | --- | --- | --- | --- | --- | --- | --- |
|  | DSC | nDSC | PPV | RVD | LTPR | LFPR | F1 |
| FLAMeS | **0.68**<sup>**††</sup> | **0.75** | **0.97**<sup>*</sup> | **0.24** | **0.75**<sup>††</sup> | **0.03**<sup>*</sup> | **0.78**<sup>**††</sup> |
| LST-AI | 0.63 | **0.75** | 0.75 | 0.25 | 0.73 | 0.24 | 0.74 |
| SAMSEG | 0.51 | 0.71 | 0.88 | 0.48 | 0.35 | 0.11 | 0.50 |

#### Cumulative Assessment

We also assessed the performance of FLAMeS vs SAMSEG when all three datasets were combined. LST methods were excluded from this analysis as LST-AI could not be applied to all datasets. Across all testing sets, FLAMeS significantly outperformed SAMSEG, with superior DSC (0.74 vs. 0.53, p<0.001), LTPR (0.87 vs. 0.37, p<0.001), and F1 scores (0.78 vs. 0.47, p<0.001), as well as higher nDSC and lower RVD (Table 6, Figure 3). FLAMeS and SAMSEG had similar PPV (0.77 vs. 0.80, p=0.46) and LFPR (0.21 vs 0.15, p=0.06).

**Table 6:**
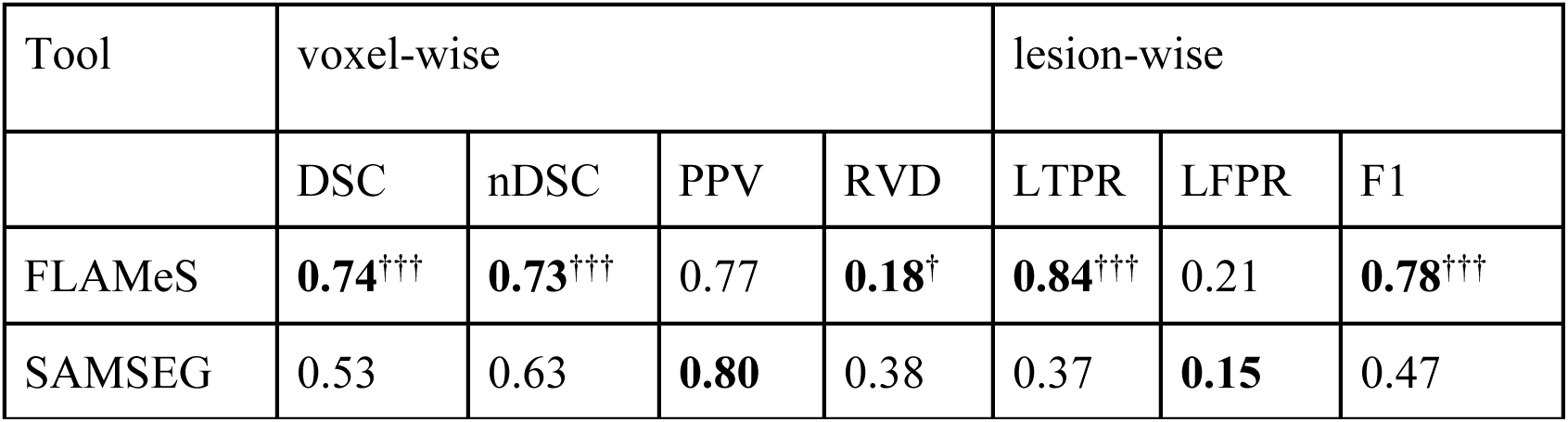
Performance comparison for FLAMeS and SAMSEG across all three testing datasets (n=75). Median values are reported for every metric. Pairwise comparisons were performed using the Wilcoxon signed-rank test. Values in bold indicate best performance for each metric. Abbreviations: DSC, Dice Similarity Coefficient; nDSC, normalized Dice Similarity Coefficient; PPV, Positive Predictive Value; RVD, Relative Volume Difference; LTPR, Lesion True Positive Rate; LFPR, Lesion False Positive Rate. Daggers indicate comparisons between FLAMeS and SAMSEG, † p<0.05, †† p<0.01, †† p<0.001

We next determined how lesion size is related to lesion detection for each method. Figure 4A depicts the lesion volume distribution across all three testing datasets. FLAMeS detects a higher proportion of smaller lesions, identifying 35% of lesions in the 3–10 mm^3^ range compared to just 6% detected by SAMSEG. Conversely, 81% of lesions over 10 mm^3^ are detected by FLAMeS compared to only 25% by SAMSEG. Since LST-AI could not be run on MSSEG-2 due to lack of T1w images in the challenge dataset, we evaluated FLAMeS against LST-AI using the remaining two datasets (Figure 4B), where FLAMeS detected 43% of lesions from 3–10 mm^3^ and 85% of lesion over 10 mm^3^, compared to 38% and 74% for LST-AI, respectively. We also found that the median volume of missed lesions was smaller for FLAMeS than for SAMSEG in every dataset as well as for LST-AI in the MSLesSeg dataset but not the clinical dataset.

**Figure 4.**
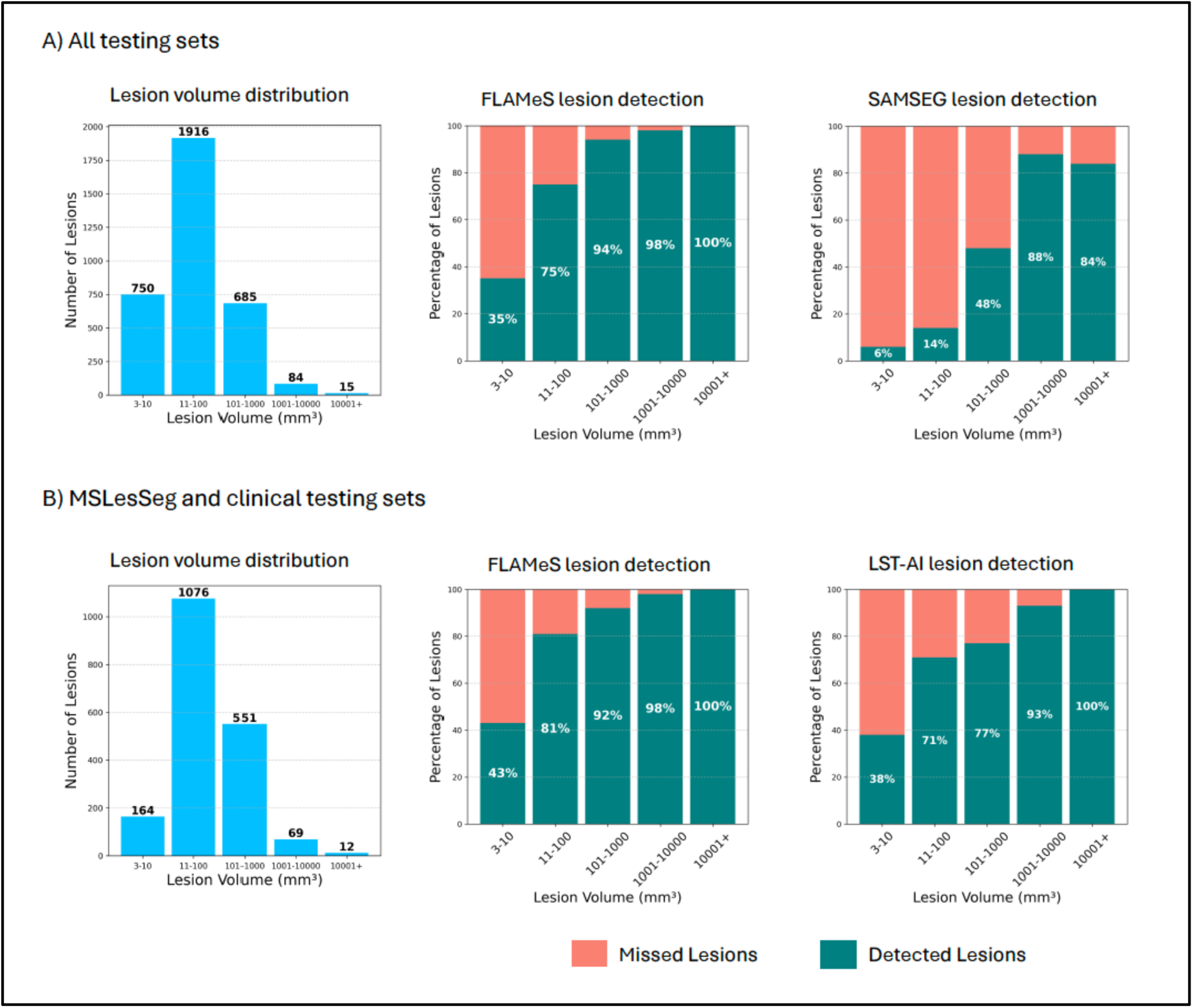
Lesion detection rates by lesion size for each segmentation method. (A) Shows the distribution of lesion volumes and detection rates by lesion size across all three testing datasets, comparing FLAMeS to SAMSEG. (B) Focuses specifically on the MSLesSeg and clinical testing datasets, showing lesion volume distribution and detection rates for these subsets, comparing FLAMeS to LST-AI.

### 3.3 Model availability

We developed a web-based version of FLAMeS, hosted on Hugging Face (Figure 5). The app features a user-friendly interface, real-time processing updates, and downloadable output files, including multiplanar snapshots of the input image and lesion mask, as well as the binary lesion mask in NIfTI format. Users also have the option to apply SynthSeg for automatic skull stripping of the input image. The app also provides an output mask with a unique label assigned to each voxel cluster to facilitate clear differentiation between lesions. Inference takes approximately one minute per MRI scan, though the app currently processes only one scan at a time due to Hugging Face’s zero-GPU usage limitations.

**Figure 5.**
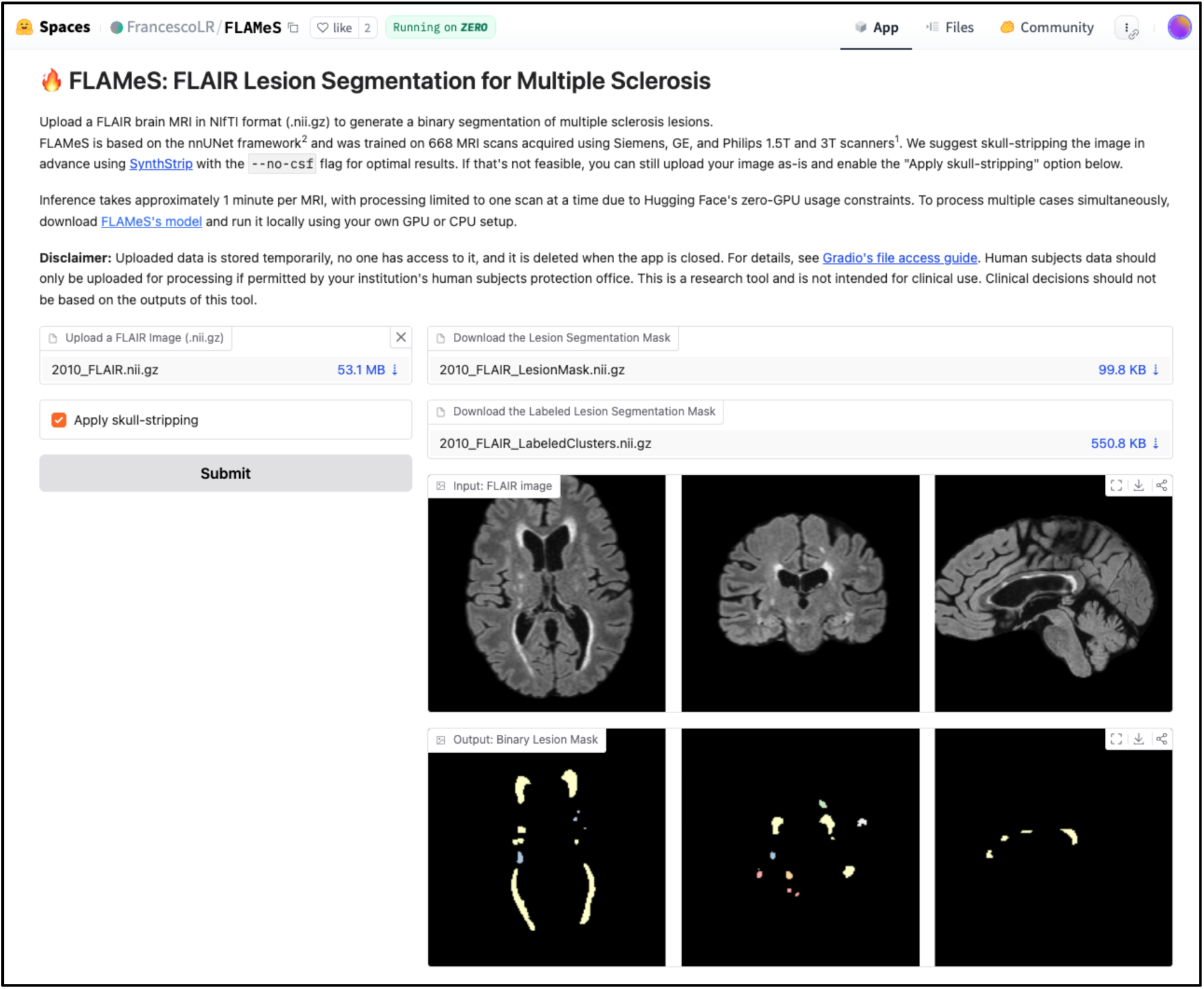
FLAMeS interactive web app.

## 4 Discussion

In this work, we propose a deep learning-based method for lesion segmentation in MS, termed FLAMeS. The model is built on a 3D nnU-Net architecture and requires only a skull-stripped FLAIR brain MRI as input. Trained on 668 FLAIR brain MRI scans from 408 individuals across seven scanning sites, the model consistently demonstrated accurate lesion segmentation across diverse acquisition parameters. To evaluate its robustness, we validated FLAMeS on three external datasets with diverse acquisition parameters. We compared its performance against two benchmark methods, SAMSEG and LST, and found that FLAMeS outperforms them across multiple metrics. These results establish FLAMeS as a reliable and widely applicable tool for automated lesion segmentation in MS. A key advantage of FLAMeS is its adaptability as it does not require retraining before being applied to new data. The only preprocessing step needed is skull-stripping, which can be efficiently performed using the SynthSeg tool from FreeSurfer.

FLAMeS delivers highly accurate lesion segmentation, effectively identifying nearly all lesions and capturing their boundaries with precision. In a blinded qualitative evaluation, two experienced raters independently selected FLAMeS segmentations as the most accurate in 15 out of 20 cases. One rater also selected FLAMeS as the top choice in two additional scans. These results highlight the accuracy, consistency, and practical utility of FLAMeS. The resulting segmentations require minimal manual adjustment and are well-suited for lesion-based analyses, especially in large datasets where substantial manual editing is often impractical.

FLAMeS consistently achieves a high DSC across all three test sets, significantly outperforming LST-AI and SAMSEG and demonstrating strong overall segmentation accuracy across diverse cases. When evaluating cumulative scores across all testing sets, FLAMeS significantly outperforms SAMSEG on every metric except for PPV and LFPR. Although FLAMeS has a marginally higher LFPR than SAMSEG, this is driven by SAMSEG’s poor lesion detection, as reflected in its very low LTPR and F1 score. In contrast, FLAMeS provides segmentations with substantially higher sensitivity and overall accuracy, as evidenced by its superior LTPR and F1 score. The slight increase in false positives may be a reasonable trade-off for robust lesion detection.

When evaluating lesion segmentation models, the F1 score is a critical metric as it reflects the balance between two key clinical priorities: accurate lesion detection (recall) and reliable prediction (precision). FLAMeS demonstrates a strong balance between these factors, achieving consistently high F1 scores and significantly outperforming LST-AI and SAMSEG across all testing sets. In addition to its high performance, FLAMeS exhibits a more favorable distribution of scores, with a tighter range across subjects and metrics (Figure 3). These findings highlight the robustness of FLAMeS at varying field strengths and on clinical and research scans.

Detecting very small MS lesions, particularly those under 10 mm^3^, remains a challenge for automated models due to their subtle appearance and the influence of the partial volume effect^28,29^. While all methods struggle with the smallest lesions, FLAMeS demonstrates greater sensitivity than LST-AI and SAMSEG (Figure 4). The lesions missed by FLAMeS tend to be smaller than those missed by the benchmark methods, indicating that FLAMeS reliably detects most larger lesions and primarily struggles with only the smallest ones. In contrast, LST-AI and SAMSEG often miss both small and large lesions, suggesting a higher threshold for consistent detection. FLAMeS’ improved sensitivity to smaller lesions enhances segmentation accuracy, reducing the need for manual corrections and ensuring a more comprehensive assessment of lesion burden.

In regard to the MSLesSeg testing set, it is important to note that this dataset differs from the other testing sets in its lesion annotation criteria. In the description of the challenge, organizers state that lesions that could not be distinctly identified on at least two consecutive slices were excluded from the ground truth lesion masks^11^. On a qualitative review, we noticed that many small lesions that our raters would have segmented were not included in the MSLesSeg ground truth masks, likely due to greater slice thickness on many of these scans (Figure 2B). This stricter annotation criteria likely accounts for the comparatively higher LFPR observed across all three methods for the MSLesSeg dataset (Table 4).

Our qualitative and quantitative assessments demonstrate that FLAMeS provides more accurate and reliable MS lesion segmentation compared to benchmark models. This prompts the question of whether FLAMeS’ superior performance stems primarily from its underlying architecture or the diversity of its training data. It is likely that both factors play a significant role. FLAMeS is built on the nnU-Net framework — a self-configuring architecture that automatically adapts to dataset-specific properties such as image size, voxel spacing, and class imbalance, eliminating the need for manual tuning^21^. Its preprocessing pipeline includes automated resampling, intensity normalization, and class balancing, making it particularly well-suited for heterogeneous datasets. In contrast, LST-AI employs an ensemble of 3D U-Nets with fixed hyperparameters and standardized preprocessing, while SAMSEG is based on a generative probabilistic framework. The difference in training data is also a key factor. FLAMeS was trained on a larger and more diverse set of 668 MS scans, compared to 491 for LST-AI and 212 for SAMSEG. Notably, LST-AI was trained exclusively on data acquired on a single 3T Achieva scanner^7^, and SAMSEG was similarly trained on data from a single site^26^. This limited diversity likely hinders the generalizability of the benchmark models to scans acquired with varied protocols, contributing to their reduced performance relative to FLAMeS.

One limitation of FLAMeS is its slight tendency toward oversegmentation, particularly in regions with intermediate signal intensity that fall between focal lesions and normal-appearing white matter (Figure 2A). A key contributor to the relative volume difference between FLAMeS segmentations and the ground truth is the differential delineation of lesion borders, influenced by partial volume effect and, most notably, these regions of intermediate-intensity, diffusely abnormal white matter (DAWM). Previous studies have reported that other automated methods also segment DAWM^30^, contributing to discrepancies between automated and manual masks, though these regions are often considered ambiguous by human raters. Oversegmentation of DAWM likely stems from the inherent limitations of a model trained solely on FLAIR images to predict lesion segmentation patterns. Notably, the benchmark methods exhibit a similar tendency toward oversegmentation (Figure 2A). To attempt to address this, we trained a modified version of FLAMeS, FLAMeS_TF, incorporating a loss function that combines Tversky and Focal losses to mitigate oversegmentation. The Tversky loss, unlike the Dice present in the standard nnU-Net loss function, allows explicit penalization of false positives. Combining it with Focal loss further emphasizes hard-to-classify voxels, possibly reducing the model’s tendency to oversegmentation. When quantitatively comparing the original FLAMeS model to FLAMeS_TF, the results are mixed. FLAMeS_TF detects significantly fewer false positive lesions. This reduction in false positives also leads to a slightly higher PPV. However, FLAMeS_TF has a lower LTPR. When considering overall performance, FLAMeS_TF achieves a similar F1 score as FLAMeS. Upon visual inspection, FLAMeS_TF produced segmentations nearly identical to the original FLAMeS model. We conclude that there is no substantial difference between the two models. Ultimately, the most effective solution may involve training the model with additional scans exhibiting DAWM.

An important next step for FLAMeS is extending its capabilities to longitudinal data, enabling precise tracking of new lesion development and change in existing lesions over time. This functionality is critical for advancing longitudinal MS research, yet remains a challenge for most existing models, particularly across heterogeneous datasets. To address this need, we plan to adapt FLAMeS for reliable and accurate lesion analysis in longitudinal data.

To conclude, we introduce an automated MS lesion segmentation tool for FLAIR brain MRI. The model requires no preprocessing beyond skull-stripping and runs efficiently, with the inference taking around 1 minute per scan. The resulting masks require little manual adjustment, in our experience taking approximately 5 to 10 minutes for the majority of cases. Our validation on three external datasets demonstrates FLAMeS’ robustness and strong performance across both research and clinical scans with diverse acquisition parameters. By offering improved accuracy and resilience to variations in image quality, resolution, and acquisition protocols, FLAMeS represents a valuable tool for MS research applications. We have made FLAMeS publicly available via a web app on Hugging Face (https://huggingface.co/spaces/FrancescoLR/FLAMeS). In addition, we have publicly released the model’s weights and code^31^ for use and to allow others to fine-tune or further improve the method.

## Data Availability

All data produced in the present study are available upon reasonable request to the authors. We also publicly release FLAMeS via web interface.

https://huggingface.co/spaces/FrancescoLR/FLAMeS

## Acknowledgments

F.L.R. received support through a Swiss National Science Foundation (SNSF) Postdoc Mobility Fellowship (P500PB_206833), Schmidt Sciences, and the Office of the Assistant Secretary of Defense for Health Affairs through the Multiple Sclerosis Research Program under Award No. (HT9425-24-1-0857). O.A. was supported by the National Multiple Sclerosis Society (FAN-1807-32163) and Department of Defense (HT9425-23-1-0571). M.W. is supported by TRAIL and the Walloon Region. J.A. acknowledges support from the NIH/NINDS (grant R01 NS131948) and NIH/NIBIB (grant P41 EB017183). Support for data collection was provided by the Intramural Research Program of the NINDS/NIH to D.S.R. (1Z1ANS003119), a grant from Bristol Myers Squibb to A.S., and NIH R01NS136523 to J.S. This work was supported in part through the computational and data resources and staff expertise provided by Scientific Computing and Data at the Icahn School of Medicine at Mount Sinai and supported by the Clinical and Translational Science Awards (CTSA) grant UL1TR004419 from the National Center for Advancing Translational Sciences. Research reported in this publication was also supported by the Office of Research Infrastructure of the National Institutes of Health under award number S10OD026880 and S10OD030463. The content is solely the responsibility of the authors and does not necessarily represent the official views of the National Institutes of Health. Opinions, interpretations, conclusions, and recommendations are those of the author and are not necessarily endorsed by the Department of Defense.

